# Quantifying the impact of contact tracing interview prioritisation strategies on disease transmission

**DOI:** 10.1101/2024.04.30.24306519

**Authors:** Logan Wu, Christopher M. Baker, Nick Tierney, Kylie Carville, Jodie McVernon, James McCaw, Nick Golding, Freya Shearer

## Abstract

Contact tracing is an important public health measure used to reduce transmission of infectious diseases. Contact tracers typically conduct telephone interviews with cases to identify contacts and direct them to quarantine, with the aim of preventing onward transmission. However, in situations where caseloads exceed the capacity of the public health system, timely interviews may not be feasible for all cases. Here we present a modelling framework for assessing the impact of different case interview prioritisation strategies on disease transmission. Our model is based on Australian contact tracing procedures and informed by contact tracing data on COVID-19 cases notified in Australia from 2020–21. Our results demonstrate that last-in-first-out strategies are more effective at reducing transmission than first-in-first-out strategies or strategies with no explicit prioritisation. To maximise the public health benefit from a given case interview capacity, public health practitioners should consider our findings when designing case interview prioritisation protocols for outbreak response.

## 1 Introduction

Contact tracing is an important non-pharmaceutical public health measure used to control infectious diseases, including COVID-19. Contact tracing aims to identify individuals (“contacts”) who may have been exposed to an infection through contact with a confirmed case during their infectious period. Once contacts are identified, public health officials may then direct them to quarantine, thereby preventing onward transmission. Contact tracing is typically a manual process where public health teams conduct detailed telephone interviews with newly confirmed cases to identify contacts.

The faster that contact tracing occurs following case exposure, the more likely that chains of disease transmission will be interrupted. However, multiple points of delay exist between the exposure of a case and identification of their contacts. These delays include, but are not limited to, the time from exposure to test, test collection to test result, test result to case notification, and case notification to case interview. The case interview step can be a significant bottleneck in contact tracing processes since interviews are time consuming, resource intensive, and limited by the number of contract tracers available [1].

When case numbers are small, as was the situation in Australia during outbreaks of COVID-19 through 2020 and early 2021, it may be possible for contact tracing teams to interview all cases on their date of notification [2]. However, when caseloads exceed the daily interview capacity, some case interviews will be delayed or missed altogether. In these situations, public health officials use various heuristics to decide which cases to interview/investigate first [1, 3]. These heuristics consider factors such as the recency of case exposure and the risk of the case transmitting to others, particularly to those at risk of severe outcomes. Risk based prioritisation can be supported by information from digital pre-interview surveys of cases [3, 4].

While the importance of risk factors for interview prioritisation is well recognised, an analysis of the impact of specific prioritisation strategies on disease transmission has not been conducted. Mathematical models of disease transmission and contact tracing processes have previously been used to quantify the impact of different contact tracing strategies on transmission of various pathogens [5–8]. While many studies have explored the impact of delays at various steps of the case and contact management process [8–11], case interview prioritisation strategies have not been investigated. Furthermore, very few modelling studies of contact tracing have considered the limited capacity of the public health system [7, 12].

Here we develop a mathematical modelling framework for assessing the impact of different case interview prioritisation strategies on disease transmission when the daily interview capacity of a public health system is exceeded. By applying our modelling framework to contact tracing data on COVID-19 cases notified in Australia from 2020–21, we estimate the reduction in transmission of SARS-CoV-2 for five case interview prioritisation strategies: oldest swab first, newest notification first, newest swab first, newest swab first then unvaccinated first within identical swab dates, and random allocation.

## 2 Methods

### 2.1 Overview

First, we develop a contact tracing queuing model that takes a series of cases as input and generates a distribution of delays from case notification to interview, according to the specified interview prioritisation strategy. The distribution of delays to interview are then fed into a stochastic model of SARS-CoV-2 transmission and contract tracing, where contacts of cases (contacts identified through case interviews) are placed in quarantine where compliance is assumed to be perfect and so no longer contribute to transmission. We then calculate the overall reduction in SARS-CoV-2 transmission due to contact tracing and quarantine for each interview prioritisation strategy. Both the queuing model and transmission model are informed by data on contact tracing delays for COVID-19 cases notified in Australia from 2020–21.

### 2.2 Data

Line-listed case data were obtained from New South Wales (NSW) Health, comprising dates of case progression through the COVID-19 case management pathway. Inputs to the queuing model include:

1. **Swab date** – when a test swab is registered at the point of collection.
2. **Confirmation delay** – time from swab collection to the health department being notified of the case/the case entering the interview queue.

### 2.3 Queuing Model

We model the interview process as a *D*^*X*^*/D*^*Y*^ */*1 queue [13], where *D*^*X*^ means that cases enter the queue as they are notified to the authority in batches of a random size *X* with a deterministic inter-arrival time of one day. To model the high variability in the number of notifications per day that we observe from the data, we model *X* as a negative binomial random variable. Cases enter the case management pathway after a delay that is randomly drawn from an empirical distribution of delays to confirmation in historical COVID-19 cases (the confirmation delay). Finally, the contact tracing team is considered as a single server with a constant daily batch capacity *Y*.

Our assumptions about the queue are that:

- The queue is not physically constrained, with unlimited capacity.
- Any cases not interviewed within five days of notification are removed from the queue (i.e., never interviewed) because extremely late interviews have little effect on transmission reduction [14]. We refer to the date five days after case notification as the ‘interview expiration date’.
- A proportion of notifications arriving on a given day cannot be interviewed on the same day due to a range of reasons such as out-of-hours notification (i.e., the interviewer or interviewee is not available) or missing contact details, even if the queue is empty. This proportion is assumed to be 20% based on the Victorian Department of Health’s COVID-19 notification system [15].
- 45% of cases are vaccinated and the vaccination status of cases is available to the contact tracing teams for interview prioritisation.

#### 2.3.1 Queue inputs

For the primary analysis, we set the mean rate for batch size, *X*, to 20 cases per day. We also examine the queuing model outputs (*i*.*e*., delay distributions) for a range of mean arrival rates where the interview capacity is a fixed proportion of the mean arrival rate (Figure A.1).

The confirmation delay for each case is drawn from empirical observations during a period of intensive contact tracing of COVID-19 cases notified in New South Wales between July 2020 and January 2021 (see Shearer and colleagues [16], Table 1).

### 2.4 Simulation

In our simulation, cases are initialised with a swab date and confirmation date. For each time step of one day, the scheduled cases enter the queue. The queue is sorted by a priority function defined by the interview prioritisation strategy, and a fixed number are processed/interviewed depending on the contact tracing capacity. Cases can be interviewed if they have not already been interviewed, they were not notified too late in the day (determined as a proportion of each day’s notifications), and their interview window has not expired.

The queue forms a discrete time Markov chain, where consecutive days are not independent (e.g., a backlog of cases from a day *d* influences who is interviewed or delayed on day *d* + 1), and the distribution of delays for a sufficiently long number of days tends to the steady-state or stationary distribution. This property of the Markov chain allows us to simulate the queue what might happen on a random day by sampling from the stationary distribution of the Markov chain. We draw sufficient samples from the stationary distribution to ensure the Monte Carlo error for the mean and standard error are negligible. A burn-in period is discarded to avoid effects from the initial empty queue where everyone is trivially processed instantly (which, for scenarios where 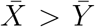, would be highly improbable in the stationary distribution). To ensure no artefacts due to right-censoring, the simulation is continued for an extra five days but any new cases entering this queue in this period are not included in the output.

#### 2.4.1 Summary of algorithm

1. On day zero, initialise the empty set of cases *C*_0_, which represents the queue of cases waiting to be interviewed.
2. For each day *t*:
  a. Generate the set of new cases *D*_*t*_ such that the number of cases |*D*_*t*_| is drawn from the random distribution ∼NB(*λ, λ*) where both mean *μ* and size *r* are equal to *λ* and so variance is *μ* + *μ*^2^*/r* = 2*λ*. These cases are ‘swabbed’ on this day and have attributes of swab date *t*, confirmation delay *δ* (the case is therefore confirmed on day *t*+*δ*, where *δ* is independently drawn from an empirical distribution), and vaccination status *v* ∈ {0, 1}, which is an independently distributed Bernoulli trial with a probability of the case being vaccinated (*i*.*e*., the proportion cases of vaccinated).
  b. Add *D*_*t*_ to the existing set of cases *C*_*t−*1_ to find the set of cases in the system on day *t*: *C*_*t−*1_ ∪ *D*_*t*_.
  c. Assign priorities to the set of current cases *C*_*t−*1_ ∪ *D*_*t*_ according to a priority function *f* (*c*) ∀*c* ∈ *C*_*t−*1_ ∪ *D*_*t*_, where *f* considers whether the case *c* is eligible for interview after the confirmation delay *δ* and vaccination status *v*. The options explored for *f* in this work are given in Section 2.5.
  d. Mark *E*_*t*_, the *n* highest-priority cases in *C*_*t−*1_ ∪ *D*_*t*_, as interviewed on day *t*, where *n* is the daily interview capacity, or the number of eligible cases, whichever is lowest. Remove *E*_*t*_ from the set *C*_*t−*1_ ∪ *D*_*t*_, such that *C*_*t*_ :=*C*_*t−*1_ ∪ *D*_*t*_ \ *E*_*t*_.
3. Increment *t* in the previous step until the total number of processed cases meets or exceeds the user-set sample size: |{*C*_*t*_ ∀*t* ∈ ℕ}| ≥ *N*_burnin_ + *N*_samples_ + *N*_burnout_
4. Discard the first *N*_burnin_ cases and use observed delay distributions from the next *N*_samples_ cases to compute the final reduction in transmission.

Note: in the implementation of this algorithm, we generate all cases in *D*_*t*_ ∀*t* with their initial attributes in an initialisation step since steps 2(a) and 3 can be determined before running the queue simulation.

### 2.5 Experimental setup

We explore five interview prioritisation strategies using the queuing model and feed outputs — the distribution of delays to interview — for each strategy into the transmission model to compute the overall reduction in transmission due to contact tracing.

1. Random swab (each day, a new sample of *n* eligible cases is interviewed)
2. Oldest swab first
3. Newest notification first
4. Newest swab first
5. Newest swab first, then unvaccinated first within identical swab dates

Furthermore, we assess each prioritisation strategy under 20%, 50%, and 80% public health workforce capacities; for example, under 20% capacity, the interview workforce is able to interview up to 20% of the mean number of incoming cases per day.

#### 2.5.1 Estimating the reduction in transmission

We estimate the reduction in SARS-CoV-2 transmission under different prioritisation strategies by feeding outputs from the queuing model (section 2.4.1) into a stochastic simulation model of SARS-CoV-2 transmission and contact tracing developed by Shearer and colleagues [16]. The stochastic simulation model is described in detail in [16]. Briefly, the model represents the relationship between contact tracing delays, symptomatic detection, and times from infection to isolation in successive chains of contact tracing. The model accounts for two modes of case detection: active detection by downstream contact tracing from the case’s infector, and passive detection by the case developing symptoms and seeking a test. By repeatedly sampling from distributions representing these processes via a recursive sampling algorithm, the model generates distributions of the time from infection to isolation for cases as described in Equations 6–13 of [16]. The distribution of times from infection to isolation are then directly translated into reductions in potential for onward transmission by following the process described by Equations 1 and 2 of [16].

The stochastic simulation model of [16] takes a number of component distributions as input, including the contact tracing delay (referred to as *T*_*C*_ in [16]). The variable *T*_*C*_ comprises several sub-component distributions including the time from swab collection to case confirmation, the time from case confirmation to case interview, and the time from interview to contact notification and swab collection. Here we supplied the model with a distribution of times for the first two of these: time from swab collection to case confirmation and time from case confirmation to case interview for each interview prioritisation strategy, as generated by the algorithm described in section 2.4.1. The time from case interview to contact notification and swab collection were as estimated by [16].

For our study, the transmission model assumes that vaccinated cases are 36% less infectious than unvaccinated cases (*i*.*e*., a 36% reduction in contagiousness given breakthrough infection). This value is based on estimates of vaccine effectiveness against onward transmission in breakthrough infections of the Delta variant of SARS-CoV-2 following two doses of the AstraZeneca (ChAdOx1 nCoV-19) vaccine as reported by Eyre and colleagues in the first version of a pre-print published in 2021 [17]. While this estimate was later updated in the peer-reviewed publication in 2022 [18], we mention it here to reflect the study conducted at the time to support Australian government decision-making on COVID-19 [19]. It should be noted that vaccine effectiveness against onward transmission in breakthrough infections is expected to be lower for Omicron variants compared to Delta [20].

The transmission model implicitly assumes that all infections are (eventually) reported as cases. In the hypothetical scenario that all cases where identified and isolated instantly after infection, this would result in a 100% reduction in transmission. However these reductions scale linearly with the rate of case ascertainment. For example, if instead only 30% of infections were detected, all of these transmission reductions would be scaled by 30%. This therefore does not influence the relative benefits of the different case interview prioritisation strategies considered here.

## 3 Results

Running our queueing simulation under different interview prioritisation strategies generates outputs of the steady-state delay distributions; i.e., a sample population of the delays experienced by a case. The different resulting distributions are shown in Figure 1. The number of missed interviews directly corresponds to the public health workforce capacity, with the number of missed interviews increasing as capacity decreases. The successful interviews are distributed from zero to five days, and the shape of these distributions depends on the prioritisation strategy. ‘Newest’ strategies exhibit shorter delays to case interview on average compared to ‘Random’ and ‘Oldest’ strategies. In Figure A.1, we demonstrate that the delay distributions from the queuing model depend only on the ratio of mean arrival rate to interview capacity, rather than the absolute value of either of these quantities. For example, for a case entering a system with 50% mean capacity, the probability of a given delay is the same irrespective of whether 10 or 1 000 cases will arrive that day (given 5 or 500 interviews respectively).

**Figure 1:**
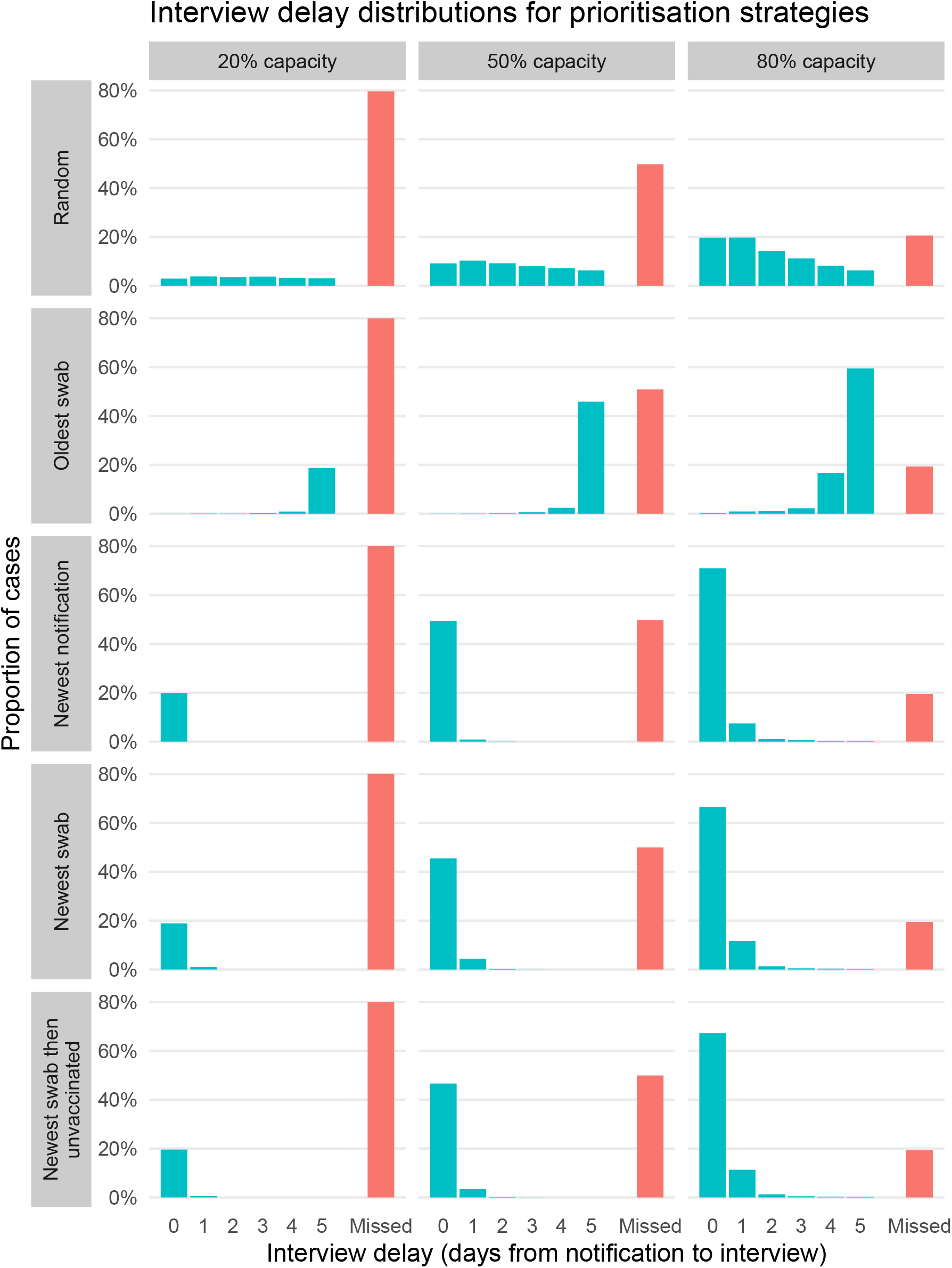
Interview delay distributions for twelve combinations of different prioritisation strategies and workforce capacities. A delay of zero days means that cases were interviewed on the same day their notification was confirmed by the health authority. If the delay exceeds five days, the interview is missed.

The delay distributions shown in Figure 1 were fed into the transmission model to estimate the overall reduction in transmission due to contact tracing, under each case interview prioritisation strategy. Outputs are shown in Figure 2 where the overall transmission reduction scales between zero (no change) and one (complete prevention of transmission). We find that the two strategies prioritising the newest swab first provide the greatest reduction in transmission, followed by newest notification first, random swabs, and finally the oldest swab first strategy. Furthermore, additional prioritisation by unvaccinated status when ordering cases swabbed on the same day provides a modest benefit in transmission reduction as vaccinated cases contribute less to downstream transmission in our transmission model.

**Figure 2:**
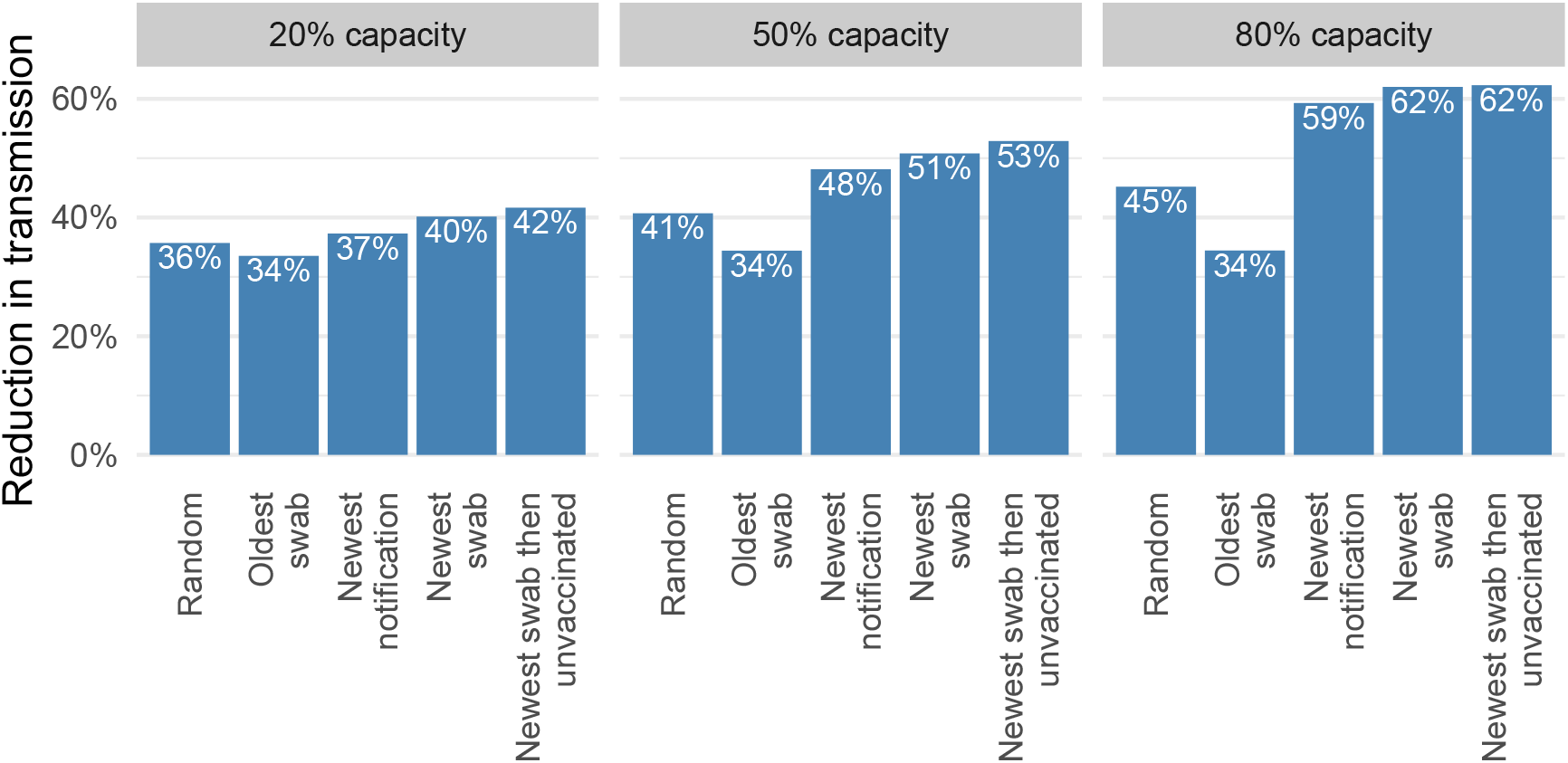
Transmission reduction factors converting the interview delay distributions using the method from Shearer et al. [16]. The simulation is run for different combinations of interview prioritisation strategy and workforce capacity as a percentage of the mean incoming case number. The reduction is the overall effect of the TTIQ system, where we assume 100% compliance with isolation and quarantine.

The ranking of strategies is consistent within a given workforce capacity, but the absolute and relative performance depends on the capacity: both the absolute transmission reduction across strategies and the difference in transmission reduction between strategies increases as the capacity increases. For example, the most effective strategy (newest swab) at 20% capacity is almost as effective as interviewing people at random in the queue at 80% workforce capacity, highlighting how an effective strategy can compensate for having far fewer interviewers. The least effective strategy (oldest swab) receives no benefit from additional capacity because the extra capacity is used to conduct interviews on the backlog of case notifications which provides little benefit in terms of transmission reduction.

## 4 Discussion

We developed a mathematical modelling framework for assessing the impact of different case interview prioritisation strategies on disease transmission and applied it to COVID-19 case data collected in Australia from 2020–21. We investigated five different case interview prioritisation strategies: oldest swab first, newest notification first, newest swab first, newest swab first then unvaccinated first within identical swab dates, and random swab.

We found that last-in-first-out strategies (newest notification first, newest swab first, and newest swab first then unvaccinated first) are more effective at reducing transmission than a first-in-first-out strategy (oldest swab first) or no explicit prioritisation (random swab). These results support an overarching principle — the case who should be interviewed next is the one where contact tracing has the potential to avert the greatest number of downstream infections. The strategy that performs best (newest swab, and its variation of prioritising vaccinated people) deprioritises those who are expected to have already contributed to the bulk of their downstream infections and, therefore, have reduced opportunity to prevent infections via contact tracing. Our results also demonstrate that the benefits of last-in-last-out strategies increase as daily interview capacity increases, *i*.*e*., the most effective strategies confer the greatest absolute and relative benefit when the daily interview capacity is highest.

A strength of our approach is in the use of data on actual contact tracing delays as input, which enabled us to generate results that were meaningful for a specific disease, population, and public health system context. Following the adoption of a national ‘re-opening’ plan in July 2021, findings from our research were reported to key SARS-CoV-2 decision-making committees in Australia in November 2021 [21] as part of a wider package of work to inform policy changes as SARS-CoV-2 became established in the broader population.

To apply our findings in practice, contact tracing teams would require support from digital infrastructure. A manually maintained spreadsheet is unlikely to be sufficient; at the very least, a programmed spreadsheet or dedicated platform is required to dynamically allocate the highest priority interview to contact tracers as each interview finishes and a new tracer becomes available.

During the COVID-19 pandemic, many countries employed smartphone-based digital contact tracing to augment manual processes, since faster and more complete contact identification is theoretically possible compared to using case interviews [22]. However, implementation issues, including poor uptake of tracing applications by smart-phone users, have largely prevented them from contributing meaningful to disease control [23]. Hence studies that aim to improve manual contact tracing processes, such as ours, remain important.

Previous mathematical modelling studies have demonstrated how reducing delays to contact tracing and/or increasing the fraction of contacts traced can reduce transmission of SARS-CoV-2 [7, 9, 11]. However, very few modelling studies of contact tracing processes have considered the limited capacity of the public health system [7, 12], and none have explored this in the context of case interview prioritisation. Kaplan and colleagues developed a model of smallpox transmission and contact tracing which, like our study, includes a tracing queue (specifically a queuing compartment in their ordinary differential equation model) where the rate at which individuals exit the queue depends upon the number of contact tracers available. However, queue exit times are not governed by an individual’s arrival time (since this was not necessary to fulfil their study aims), and so would be equivalent to our random swab strategy [12]. Meister and Kleinberg developed an algorithm for determining the optimal ordering for tracing of identified contacts, according to each contact’s probability of infection, recency of exposure, and the number of other contacts they may have exposed [24]. However, they do not model the impact of contact tracing on transmission as they only consider the direct benefits of timely medical treatment due to contact notification. Indirect clinical benefits arising from the prevention of onward transmission via contact tracing and quarantine are therefore not considered.

Our study has several limitations. Since case data were provided at a daily resolution, it was necessary to process simulations in daily batches so that empirical delays could be incorporated in our model without making intra-day assumptions. While this is more computationally efficient than sub-daily processing, if data were available at a sub-daily resolution (*e*.*g*., with date stamps), it would be possible to explore additional processing considerations such as the relative merits of batch processing versus online processing (where the queue order would be updated in real-time in response to newly notified cases). For pathogens such as SARS-CoV-2 where the contact tracing window is relatively short and small reductions in delays can lead to significant impact on disease control [9], shorter-than-daily batch processing or online processing may result in further reductions in transmission.

Our study does not consider risk factors for onward transmission except for the vaccination status of cases (since we assume that unvaccinated cases are more infectious than vaccinated cases). Other factors such as case occupation, age, or housing type, or whether a case visited or worked in a high-risk location/setting during their infectious period, are often incorporated in protocols for case interview prioritisation [1, 4, 14, 25]. For example, contact tracers may prioritise interviewing a person who works in a healthcare setting over an academic working from home, since the healthcare worker likely has many more in-person interactions each day than the academic. In many countries, interviews of COVID-19 cases linked to high-priority settings such as aged care were expedited by health departments [14, 25]. A more comprehensive risk prioritisation model would assume the existence of pre-interview case surveys, potentially conducted at the time of swab collection, so that information on key risk factors is available to a prioritisation algorithm as soon as newly confirmed cases enter the interview queue. The relationship between these variables and onward transmission would also need to be quantified to make full use of them within a prioritisation algorithm. Nonetheless, if these additional variables were incorporated in a prioritisation algorithm, we expect that our overarching findings would hold, because any delay to interview for these ‘high risk’ cases would result in less benefit in terms of disease control. Finally, our prioritisation algorithm only uses vaccination as a ‘tiebreaker’ for cases with the same swab/notification date. Other prioritisation strategies could be explored where the relative onward transmission risk for cases exhibiting different combinations of risk factors is considered.

Our findings can inform outbreak response for COVID-19 and other diseases with similar biological characteristics affecting transmission. Contact tracing is time-sensitive for any disease that spreads slowly enough to enable a tracing window but sufficiently fast that even small reductions in tracing delays can have a large benefit for outbreak control [5]. While the exact numerical results provided here will vary depending upon the infectious disease and the population in which the disease is spreading, our modelling framework can be applied to assess the impact of interview prioritisation strategies using contact tracing data collected in other contexts. Furthermore, our approach is applicable to other case and contact management processes that are both time-sensitive and where public health workloads may exceed the system capacity, including diagnostic testing procedures.

## Data Availability

All code provided to simulate the data are provided online in the Github repository

https://github.com/loganbwu/ttiq-queueing

## 5 Code availability

All analyses are performed in R. Code is available at https://github.com/loganbwu/ttiq-queueing.

## 6 Data availability

The analyses performed in this study required access to data from the New South Wales (NSW) Notifiable Conditions Information Management System (NCIMS), which are not able to be shared under the terms of our data access agreements. For access to NCIMS data, contact the NSW Ministry of Health via https://data.nsw.gov.au/contact.

## 7 Ethics statement

The study used routinely collected patient administration data from the New South Wales (NSW) Notifiable Conditions Information Management System (NCIMS). De-identified NCIMS data were securely managed to ensure patient privacy and to ensure the study’s compliance with the National Health and Medical Research Council’s Ethical Considerations in Quality Assurance and Evaluation Activities (https://www.nhmrc.gov.au/about-us/resources/ethical-considerations-quality-assurance-and-evaluation-activities). These data were provided for use in this study to support public health response under the governance of Health Protection NSW. The NSW Public Health Act (2010) allows for such release of data to identify and monitor risk factors for diseases and conditions that have a substantial adverse impact on the population and to improve service delivery. Following review, the NSW Ministry of Health determined that this study met that threshold and therefore provided approval for the study to proceed. The project oversight and approval for publication was provided by the NSW Ministry of Health.

## 8 Acknowledgements

This work was directly funded by the Australian Government Department of Health Office of Health Protection. Additional support was provided by the National Health and Medical Research Council of Australia through its Centres of Research Excellence (SPECTRUM, GNT1170960) and Investigator Grant Schemes (JMcV Principal Research Fellowship, GNT1117140; FMS Emerging Leader Fellowship, 2021/GNT2010051).

This research was supported by The University of Melbourne’s Research Computing Services and the Petascale Campus Initiative.

This work could not have been completed without the support of the public health network in NSW. The Public Health Response Branch, NSW Health including contact tracing staff as well as operational, policy, epidemiology, surveillance, data quality and data acquisition staff contributed to the TTIQ implementation and data provision. In addition, colleagues in the public health units located within the 15 local health districts across the state all contributed to the implementation of the TTIQ strategy.

## A Mean arrival rate of the queuing model

**Figure A.1:**
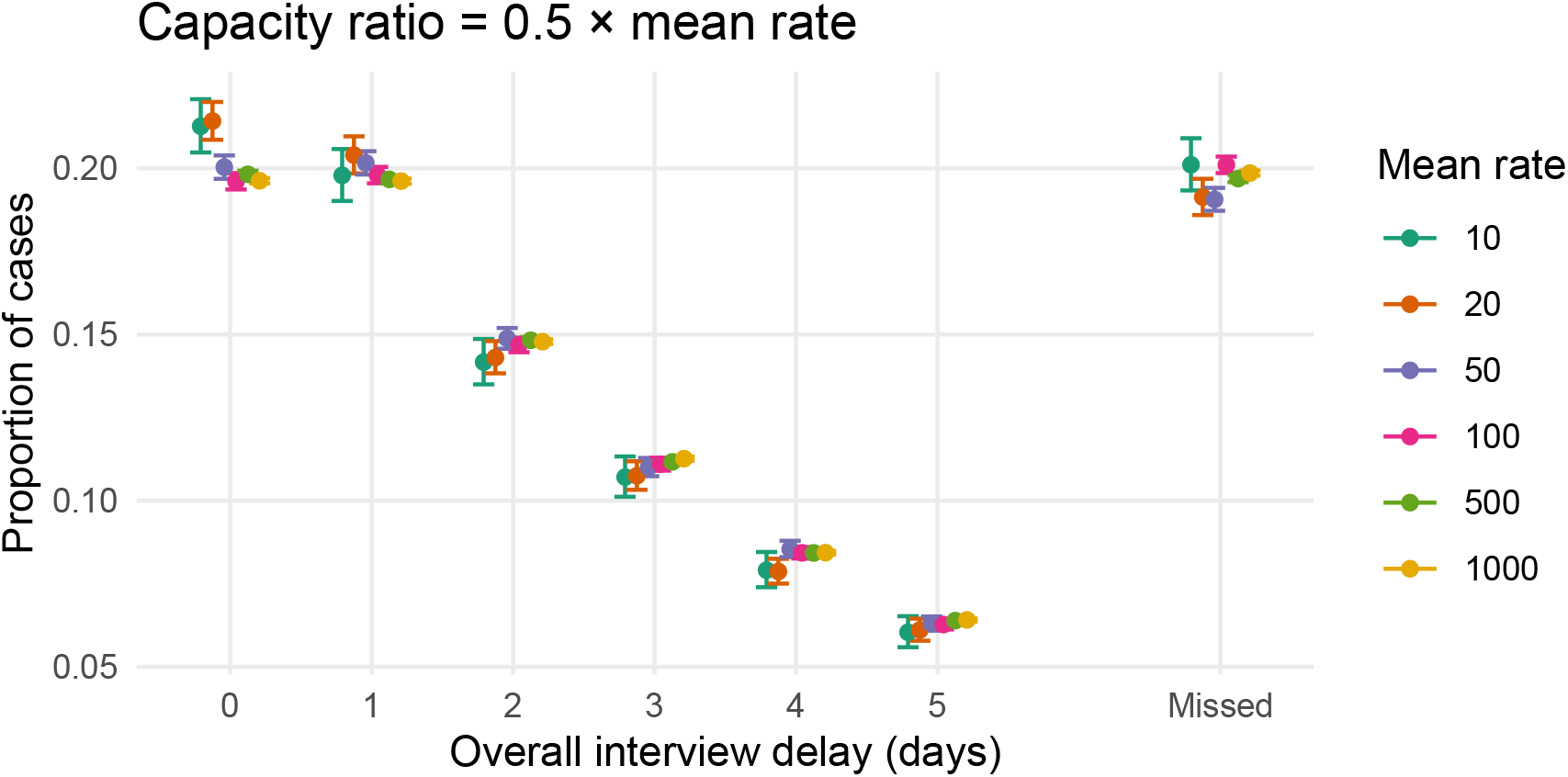
Interview delay distributions (*i*.*e*., queuing model outputs) under the ‘random’ interview prioritisation strategy exhibit similar mean values (dots) for a range of mean arrival rates where the interview capacity (50%) is a fixed proportion of the mean arrival rate of cases into the queue. Intervals show variation of the distribution after simulating 1 000 days, represented as a 95% Wilson CI of the mean proportions.

